# Brain Network Excitability Predicts Clinical Severity in Multiple Sclerosis

**DOI:** 10.64898/2026.07.10.26357763

**Authors:** L.G. Amato, M. Angiolelli, M. Demuru, E. Troisi Lopez, M. Quarantelli, C. Granata, D. Depannemaecker, V. Jirsa, S. Bonavita, A. Mazzoni, P. Sorrentino

## Abstract

Comprehensive biomarkers of multiple sclerosis (MS) capable of simultaneously diagnosing the condition, capturing symptom severity and predicting treatment efficacy remain elusive. Although several studies have highlighted the pivotal role played by demyelinating lesions in determining MS structural pathology, their relationship with symptom severity is limited. Here, we combined personalized computational brain modeling with magnetoencephalography (MEG) recordings from 17 MS patients and 20 healthy controls (CTR) to derive personalized brain network excitability parameters, which we tested as MS biomarkers. Personalized parameters discriminated between CTR and MS participants with high accuracy, also classifying between progressing and remitting MS patients. Notably, they also predicted MS clinical scales across multiple domains. In all clinical tasks, personalized parameters consistently outperformed standard clinical measures and total lesion loads. Together, these results highlight the potential of personalized brain modelling in deriving integrative MS biomarkers, capable of simultaneously identifying the condition, classifying MS subtypes and predicting symptom severity.

## Introduction

Multiple sclerosis^1^ (MS) is a neurodegenerative disorder characterized by marked clinical heterogeneity and a complex relationship between structural damage and functional impairment^2,3^. Despite significant advances in neuroimaging techniques, the identification of reliable quantitative biomarkers to univocally diagnose the condition and predict disability remains an open challenge^4^. In particular, total white matter (WM) lesion load, historically used as an index of disease severity, has yielded inconsistent results in predicting clinical progression^2,5^.

Multiple sclerosis has traditionally been conceptualized as a disease of inflammatory demyelination^6^, and this view has strongly shaped computational modeling approaches to the disorder. In particular, previous modeling studies have often focused on alterations in conduction delays, under the assumption that myelin damage primarily affects the timing of signal propagation along structural connections^7^. This strategy is well suited to capture one central aspect of MS pathophysiology, namely the disruption of white-matter transmission. However, demyelination and altered conduction velocity represent only part of the disease process, and do not fully account for the broad spectrum of clinical disability, disease progression, and neurodegenerative changes observed in patients^2^.

Increasing evidence indicates that gray matter involvement plays a crucial role in MS, both clinically and biologically^8^. Cortical and deep gray matter damage are now recognized as major contributors to disability, cognitive impairment, and disease progression^9^. This is supported by imaging studies showing that sequences sensitive to cortical lesions, such as double inversion recovery imaging, reveal gray matter abnormalities that are strongly associated with clinical disability^10^. Thus, focusing exclusively on conduction delays may overlook a complementary and potentially critical dimension of MS pathophysiology: altered local excitability within gray matter regions.

This issue is particularly relevant in secondary progressive MS, where neurodegenerative mechanisms become increasingly prominent relative to acute inflammatory demyelination^11^. In this stage, clinical worsening may reflect not only impaired communication between regions due to myelin damage, but also changes in the intrinsic dynamical properties of cortical and subcortical gray matter, which may serve as a better indicator of future disease progression^12^.

Several functional^13^ and neurophysiological^14,15^ metrics to quantify and predict the functional effect of the structural involvement have been proposed, yet robust, objective, and sensitive quantitative measures are still lacking^16^. Particularly, multiple alterations in excitability have been related to MS pathophysiology^17–19^. Recent studies have suggested that computational modeling may offer a promising avenue to bridge this gap^20–23^. In this context, mechanistic whole-brain models should not be viewed as purely descriptive or correlative tools, but as formalized hypotheses about how local neural dynamics, anatomical connectivity, and biophysical parameters interact to generate the brain activity observed empirically. In epistemic terms, they provide an intermediate level of explanation between macroscopic imaging markers and microscopic pathophysiological mechanisms: rather than asking only whether a given imaging feature correlates with disability, they ask which latent physiological changes must be assumed for the model to reproduce the observed brain dynamics. Thus, model parameters can be interpreted as candidate mechanistic variables, offering a way to infer otherwise unobservable properties of the diseased brain from measurable structural and functional data.

In previous works^20,21^, brain models have been shown to predict clinical disability, as measured by the Expanded Disability Status Scale (EDSS), by estimating subject-specific parameters related to neural signal propagation through simulation-based inference approaches. These models provide a mechanistic framework for linking structural connectivity to emergent brain dynamics, enabling the extraction of latent physiological parameters that are otherwise inaccessible from empirical data alone.

In this work, we applied a digital twin framework to 37 participants, 17 MS and 20 healthy controls (CTR, Fig 1a.i). Among MS participants, 7 were relapsing-remitting MS (RRMS) and 10 were secondary progressive MS (SPMS). All participants underwent structural MRI (Fig 1a.ii) and MEG recordings (Fig. 1a.iii). We applied personalized computational brain modeling to reconstruct excitability-related metrics, including synaptic velocity and background activity of different neuronal populations in both healthy controls and MS patients. These excitability metrics are hypothesized to be altered by the MS pathophysiology^17–19^. Individualized models (digital twins, Fig 1b.i) were built using subject-specific structural connectivity derived from diffusion MRI tractography, while magnetoencephalography (MEG) recordings were used as functional data to constrain and validate simulated dynamics. Through this multimodal integration, a genetic algorithm was used to reconstruct subject-specific alterations in neural excitability associated with MS, which were then tested as computational biomarkers (Fig 1b.ii).

**Figure 1:**
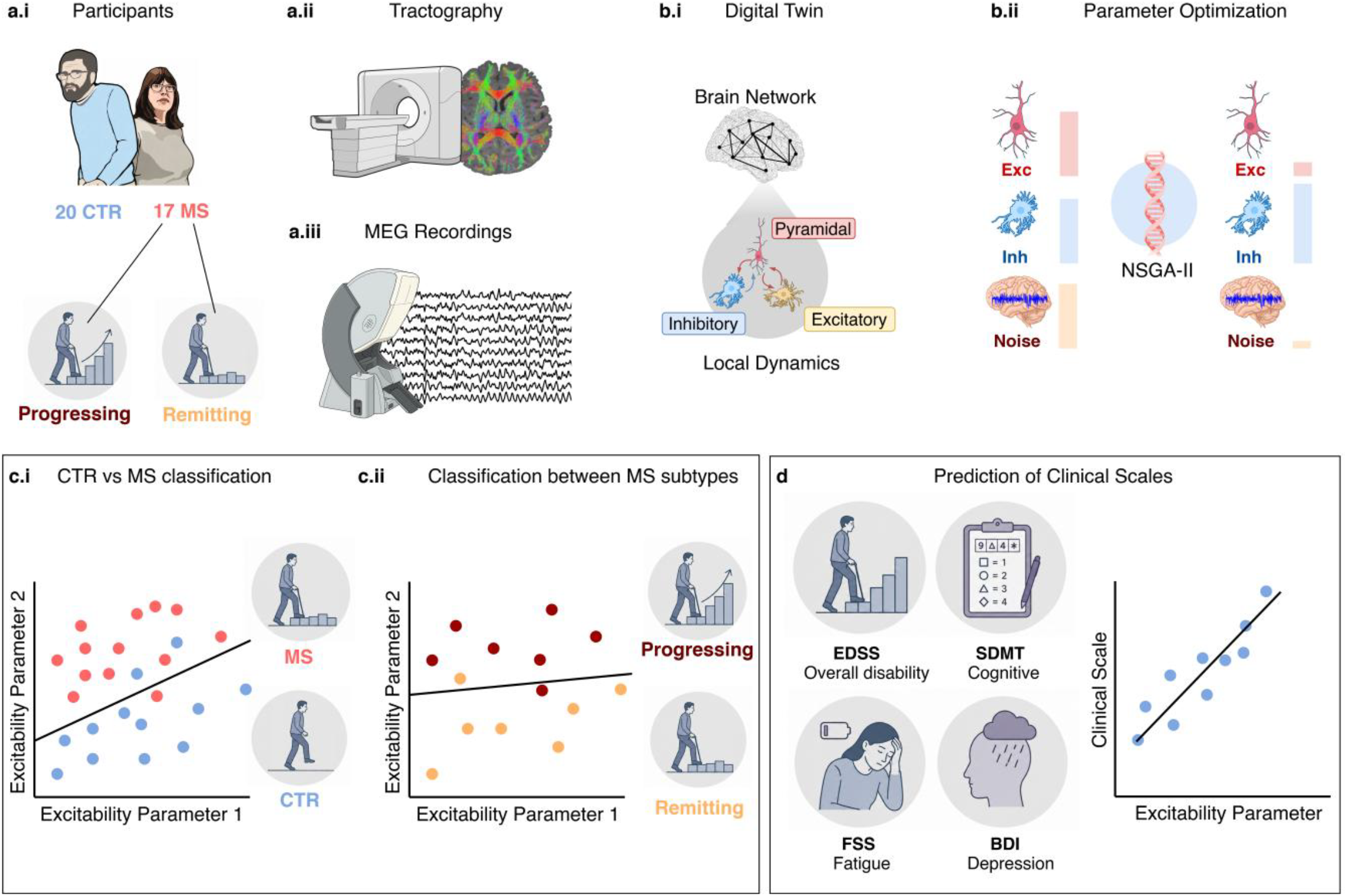
Computational workflow. **(a):** 37 participants (i), 20 of which CTR and 17 MS, (of whom 7 of the RRMS and 10 of the SPMS subtype) underwent structural MRI to extract personalized tractography data (ii) and MEG recordings (iii) to sample neural activity. **(b):** Digital twins were reconstructed for each participant (i), optimizing individual parameters with the NSGA-II genetic algorithm (ii). **(c):** Personalized excitability parameters were tested to classify between CTR and MS participants (i) and between patients with different MS subtypes (ii). **(d):** The association of excitability parameters with MS clinical scales was tested.

Our results show that excitability parameters robustly discriminate between healthy controls and MS patients, and further support the classification of both CTR vs. MS participants (Fig. 1c.i) and between relapsing-remitting and progressive MS subtypes^24^ (Fig. 1c.ii). Importantly, these computational markers also exhibit significant associations with clinical and cognitive scales (Fig. 1d), including EDSS^25^, Beck Depression Inventory^26^ (BDI), Fatigue Severity Scale^27^ (FSS), and Symbol Digit Modalities Test^28^ (SDMT).

Overall, our findings support the potential of personalized computational models as powerful tools for deriving mechanistic, quantitative biomarkers of disease severity in MS. Importantly, this work extends the virtual-brain-twin framework in MS beyond assessing demyelination-related alterations in signal propagation, highlighting its ability to capture neurodegenerative mechanisms associated with gray matter dysfunction. In this perspective, personalized brain modeling may provide a broader mechanistic account of MS pathophysiology, linking structural damage, altered brain dynamics, and clinical outcomes beyond a purely autoimmune-inflammatory or white-matter-centered view of the disease.

## Methods

### Participants, MEG and MRI recordings

Participants’ recruitment, alongside the specifications of MEG recordings and preprocessing is reported in Sorrentino et al.^20^ The tractography and lesion load derivation from MRI recordings can be consulted in Sorrentino et al.^20^

Briefly, participants with multiple sclerosis were recruited from the outpatient clinic of the Institute for Diagnosis and Cure Hermitage Capodimonte in Naples, Italy. Diagnosis was established according to the revised 2017 McDonald criteria^29^, and patients were excluded in the presence of recent relapse or steroid therapy, relevant neurological or psychiatric comorbidities, substance use, inability to complete clinical/cognitive assessments, or contraindications to MRI. All patients underwent standardized neurological and neuropsychological assessments, including the Expanded Disability Status Scale score, the Symbol Digit Modalities Test, the Fatigue Severity Scale, and the Beck Depression Inventory. Healthy controls were recruited among non-genetically related caregivers and spouses. The study was conducted in accordance with the Declaration of Helsinki and approved by the local Ethics Committee, with written informed consent obtained from all participants.

MRI and MEG data were acquired and processed as reported previously. In brief, MRI acquisition included high-resolution T1-weighted, FLAIR, and diffusion-weighted sequences, allowing quantification of white matter lesion burden, global atrophy, and reconstruction of subject-specific structural connectivity. White matter lesions were segmented from FLAIR images and lesion filling was applied before tissue segmentation to reduce lesion-related bias. Diffusion MRI data were corrected for motion and eddy-current distortions, and whole-brain deterministic tractography was used to reconstruct anatomical connectivity between cortical regions defined according to the Desikan-Killiany-Tourville atlas. Resting-state MEG recordings were preprocessed to remove noisy channels, environmental noise, and physiological artifacts, including cardiac and ocular components. Source-level activity was then reconstructed for 84 cortical regions using a Linearly Constrained Minimum Variance beamformer based on each participant’s anatomical MRI. These multimodal data provided the empirical and anatomical basis for the subsequent personalized whole-brain modeling analyses performed in the present study.

### Parameter derivation via multi-objective optimization

Subject-specific model parameters were estimated using a multi-objective evolutionary optimization framework based on the NSGA-II genetic algorithm^30^. For each participant, we instantiated a whole-brain network model constrained by individual structural connectivity matrices derived from diffusion data. The computational model was developed using The Virtual Brain^31,32^ software, using the Jansen-Rit model^33^ to simulate local neural activity of brain regions.

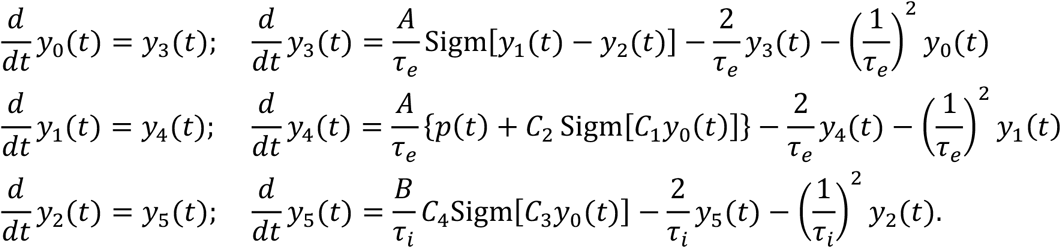

The parameters 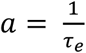 and 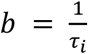 represent the inverse of (respectively) excitatory and inhibitory time constants, and were considered among the excitability parameters affected by the disease (and thus reconstructed with the genetic algorithm). External neural activity is injected into each local node by the term *p*(*t*) = *µ*(*t*) + *σ*(*t*), where *µ*(*t*) represents the input coming from other nodes and *σ*(*t*) represents background activity (neural noise). For each region *j*, the term *µ*(*t*) is modelled as the sum of activity of other *i* regions *ω*_*i*_, which is injected into *j* with a value *µ*(*t*) = *g* ∑_*i*_ *c*_*i,j*_*ω*_*i*_(*t* − τ_*D*_) depending on:

- The connection between *i* and *j*: *g* × *c*_*i,j*_, where the global connectivity constant *g* was reconstructed for each patient with the genetic algorithm, and *c*_*i,j*_ from the structural connectivity (*SC*) matrix obtained from individual tractography data of each patient.
- The delay time 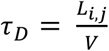, given by the ratio between the physical distance between nodes *L*_*i,j*_ derived for each patient from individual tractography data and the conduction velocity constant *V*, reconstructed for each patient with the genetic algorithm.

The *σ*(*t*) term was modelled as additive Gaussian white noise, whose amplitude was reconstructed for each patient with the genetic algorithm. The model included a set of biophysically interpretable parameters governing large-scale dynamics: global connectivity coupling (g), determining the strength of the connective weights of the structural connectivity (*SC*) matrix; conduction velocity, stochastic noise amplitude (background activity), and local dynamical parameters reflecting the velocity of excitatory and inhibitory synapses (a, b). For MS patients, an additional parameter (here referred to as *sevcut*) was introduced to capture the impact of lesion burden on network communication. Lesion maps (*LM*) were incorporated to inform this *sevcut* parameter, which was subtracted from the structural connectivity to determine connective weights *c*_*i,j*_ with the equation:

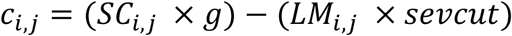

Specifically, lesion masks were used to quantify both the total lesion load and its distribution across the connectome. This information was integrated into the model by modulating the strength of “effective” structural connectivity, allowing the optimization procedure to capture subject-specific structural damage beyond what is reflected in the tractography-derived weights alone.

The simulations were conducted to generate 50 s of resting-state activity in each region of the implemented parcellation with a uniform set of parameters. For each simulation, PSD and FC features were extracted following the same procedure adopted for empirical data.

Model inversion was performed by iteratively simulating neural activity and comparing simulated outputs with empirical neurophysiological data. Two complementary empirical targets were used: (i) functional connectivity (FC) computed in the alpha band (8 – 13 Hz), and (ii) regional power spectral density (PSD) in the 0.5 – 13 Hz range. Empirical features were selected in this range since the JR model is meant to simulate activity from low frequencies to high alpha frequency (0 – 13 Hz). While PSD was simulated in this whole range, FC -which is usually divided into frequency bands-was simulated in the alpha range. This was based on a preemptive mutual information analysis of FC bands which selected the alpha band as the most informative (*MI*_*alpha*_ = 0.098, *MI*_*theta*_ = 0.067, *MI*_*delta*_ = 0.001). The optimization aimed to minimize the discrepancy between simulated and empirical FC (quantified as mean absolute error) and between simulated and empirical PSD profiles. These two objectives were jointly optimized using NSGA-II, yielding for each subject a Pareto front of optimal solutions representing trade-offs between the two metrics. For each participant, 20 generations of 10 individuals each were generated. From this front, representative parameter sets were selected for subsequent analyses.

The resulting parameters can be interpreted as digital biomarkers reflecting large-scale brain dynamics. From the primary parameters (global connectivity coupling, conduction velocity, background activity, and the velocity of excitatory and inhibitory synapses) we derived additional parameters related to both structural connectivity and excitability features. These included: (i) node degree, (ii) ratios between excitatory and inhibitory parameters (a/b), (iii) connectivity coupling over cut intensity.

Moreover, regional values of parameters were computed to investigate the interplay between excitability parameters and network topology^34,35^. These quantities (excitability parameters weighted by node degree) were computed by multiplying excitability parameters (uniform in all brain regions) with the node degree of different subnetworks. This allowed for the computation of excitability parameters not only at the whole-brain level but also within anatomically defined subnetworks. These included canonical anatomical partitions (frontal, temporal, parietal, occipital lobes and hemispheric divisions). This multiscale characterization was chosen to probe region-specific alterations in brain dynamics and to identify network-level signatures of disease.

### Classification analyses (CTR vs. MS and RRMS vs. SPMS)

To identify the most informative biomarkers, we systematically evaluated excitability parameters based on their ability to discriminate between CTR and MS participants. The non-parametric Mann-Whitney U-test was used to assess differences in median values across groups, and candidate biomarkers were ranked based on effect size and significance.

In parallel, we evaluated their discriminative power using balanced accuracy as a performance metric (simply referred as “accuracy” from now on for the sake of brevity). Briefly, the selected biomarkers were then used as features in binary classification tasks. Only linear classifiers were considered (support vector machines with linear kernel) with a maximum of two classifying features, to restrict our analysis to strong and interpretable decision boundaries relating biomarker values with disease status. Receiver operating characteristic (ROC) curves and confusion matrices were used to quantify classification performance. Decision boundaries were explicitly visualized at each classification to provide an interpretable representation of class separability. Due to the low sample size and the choice of restricting to linear and interpretable decision boundaries, no cross-validation was performed on the classifications.

The same framework was extended to stratify MS patients into clinical subtypes, namely, relapsing-remitting MS and secondary progressive MS. Biomarkers identified at the group level were used to construct decision boundaries in the reduced feature space, enabling visualization and quantification of subtype separability. This approach allowed us to assess whether the derived dynamical biomarkers capture clinically meaningful heterogeneity within the MS population.

For classification of MS subtypes, standard clinical features (age, sex, education level, disease duration, and total lesion load from MRI scans) were included alongside excitability parameters as classification features.

### Prediction of clinical MS scales

To investigate the clinical relevance of the derived biomarkers, we modeled their relationship with established clinical scales of disease severity and symptomatology (EDSS, FSS, SDMT, BDI). For each clinical scale, we constructed multivariate regression models using combinations of excitability parameters as predictors. Performance was benchmarked against that of a multivariate regression based on standard clinical features (see previous paragraph). To establish the added value of excitability parameters over standard neurophysiological analysis, we also assessed the performance of a multivariate regression model based on combined standard MEG features (functional connectivity and power spectral density) and clinical features

Given the small sample size, model validation was performed using a leave-one-out cross-validation (LOO-CV) scheme. In this framework, each subject was iteratively excluded from the dataset, the model was trained on the remaining subjects, and predictions were generated for the held-out individual. This procedure was repeated for all subjects, and predictive performance was quantified by averaging the coefficient of determination (R^2^) across folds. This approach provides an unbiased estimate of model generalizability while maximizing the use of available data.

Feature importance was assessed using permutation-based methods^36^, for each prediction. In addition, we evaluated the stability of model performance by assessing variability across folds. This framework enabled us to link mechanistic model parameters with large-scale data features and, in turn, clinically meaningful outcomes, providing insights into how large-scale brain dynamics relate to symptom severity and disease progression.

### Statistical Analysis

Group differences between CTR and MS participants were assessed using non-parametric Mann-Whitney U tests. Categorical variables (sex) were compared using chi-square (*χ*^2^) tests. Effect sizes were quantified using Cohen’s d. In classification tasks, performance was quantified using metrics such as accuracy, area under the ROC curve (AUC), and confusion matrices. Statistical thresholds were set a priori, and results were interpreted alongside effect sizes to ensure robustness and practical relevance. For clinical scales prediction, multiple regression analyses were conducted to identify aggregate predictors. For each clinical scale, we computed the R^2^ coefficient and adjusted estimates using leave-one-out cross-validation. The correlation between clinical scales and single biomarkers was assessed using Pearson’s correlation.

## Results

We analyzed resting-state MEG recordings collected from 20 CTR and 17 MS participants. No statistically significant differences were found across groups in term of demographics (Table 1A). Among MS participants,10 were of the SPMS type and 7 of the RRMS subtype. Across MS subtypes, no differences were found in demographics, clinical scales and total lesion load (Table 1B), with the exception of EDSS differences between SPMS and RRMS patients, which did not survive multiple-comparison correction (U = 53.5, p = 0.021).

**Table 1:**
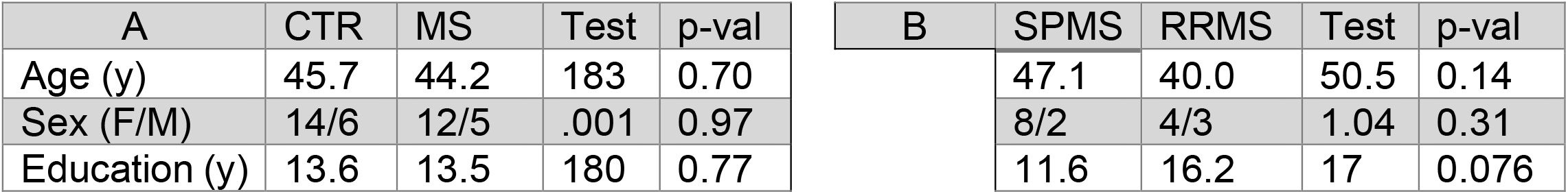
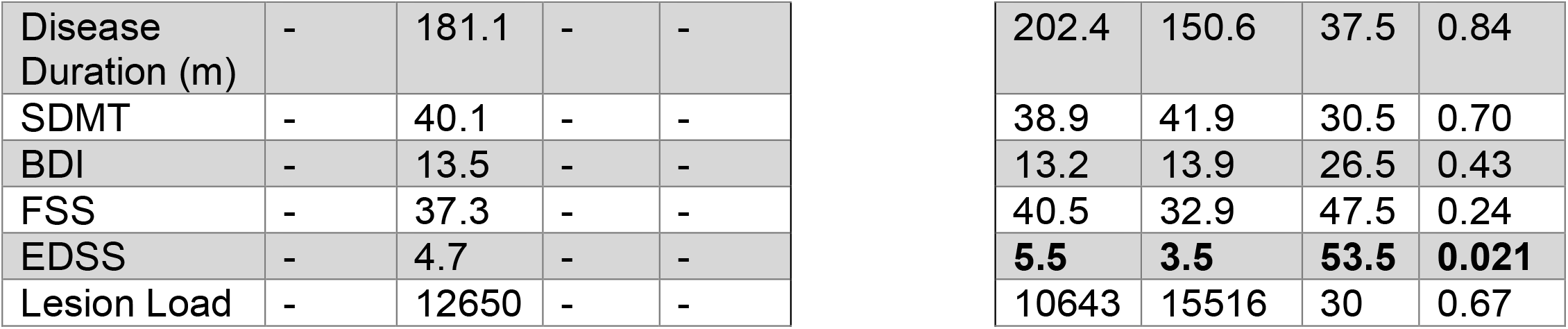
Demographic and clinical information of enrolled participants. **(A):** Mean values in CTR and MS participants. For biological sex, number of females and males is reported. Statistical test values and p-values of the Mann-Whitney U-test are also reported. **(B):** Mean values in MS subtypes (SPMS and RRMS). Notation is the same as in (A). Significant differences (prior to multiple comparisons correction) are highlighted in bold.

| A | CTR | MS | Test | p-val |
| --- | --- | --- | --- | --- |
| Age (y) | 45.7 | 44.2 | 183 | 0.70 |
| Sex (F/M) | 14/6 | 12/5 | .001 | 0.97 |
| Education (y) | 13.6 | 13.5 | 180 | 0.77 |

| B | SPMS | RRMS | Test | p-val |
| --- | --- | --- | --- | --- |
|  | 47.1 | 40.0 | 50.5 | 0.14 |
|  | 8/2 | 4/3 | 1.04 | 0.31 |
|  | 11.6 | 16.2 | 17 | 0.076 |

**Table 1: Demographic and clinical information of enrolled participants.**
|  |  |  |  |  |  |  |  |  |
| --- | --- | --- | --- | --- | --- | --- | --- | --- |
| Disease Duration (m) | - | 181.1 | - | - | 202.4 | 150.6 | 37.5 | 0.84 |
| SDMT | - | 40.1 | - | - | 38.9 | 41.9 | 30.5 | 0.70 |
| BDI | - | 13.5 | - | - | 13.2 | 13.9 | 26.5 | 0.43 |
| FSS | - | 37.3 | - | - | 40.5 | 32.9 | 47.5 | 0.24 |
| EDSS | - | 4.7 | - | - | **5.5** | **3.5** | **53.5** | **0.021** |
| Lesion Load | - | 12650 | - | - | 10643 | 15516 | 30 | 0.67 |

### Excitability parameters discriminate between CTR and MS patients and between MS subtypes

No significant differences were observed between the CTR and MS groups in terms of sex (*χ*^2^ = 0.001, p = 0.97), education (U = 180, p = 0.77), or age (U = 183, p = 0.70), indicating good matching between groups. Similarly, SPMS and RRMS subgroups did not differ significantly in disease duration (U = 37.5, p = 0.84), sex (*χ*^2^ = 1.04, p = 0.31), lesion load (U = 30.0, p = 0.67), education (U = 17.0, p = 0.076), or age (U = 50.5, p = 0.14).

Subject-specific model parameters were estimated for all participants using a simulation-based inversion procedure constrained by PSD and FC derived from MEG recordings. The inferred parameters included total global connectivity coupling, conduction velocity, excitatory synaptic velocity, inhibitory synaptic velocity, and background activity. Among these parameters, only background activity revealed statistically significant differences between MS and CTR after Bonferroni correction: Global connectivity coupling: U = 259, p = 0.022, d = 0.64, conduction velocity: U = 181, p = 0.98, d = -0.03, Global background activity: U = 309, p = 0.0002, d = 1.14, excitatory synaptic velocity: U = 242, p = 0.072, d = 0.38, inhibitory synaptic velocity: U = 69, p = 0.013, d = -0.42.

In addition to these primary parameters, composite metrics were computed by combining the original parameters and by weighing them within anatomically defined subnetworks (submatrices of the structural connectivity matrix corresponding to frontal, temporal, parietal and occipital regions, and left/right hemispheres). This was designed to measure the interaction between excitability and network topology (see Methods).

An automated feature selection procedure identified the parameters that best discriminated between CTR and MS, including primary and composite parameters. The most informative features were the global weighted background activity (U = 313, p = 0.0001, d = 1.51) and the excitation-to-inhibition balance, defined as the ratio between excitatory and inhibitory synaptic velocities (U = 287, p = 0.002, d = 1.24). Both parameters showed large effect sizes, indicating strong group separation.

These two features were further combined to define a two-dimensional decision boundary (Fig. 2a), achieving high classification performance between CTR and MS, with an AUC of 0.87 and an accuracy of 0.87 (Fig. 2b).

**Figure 2:**
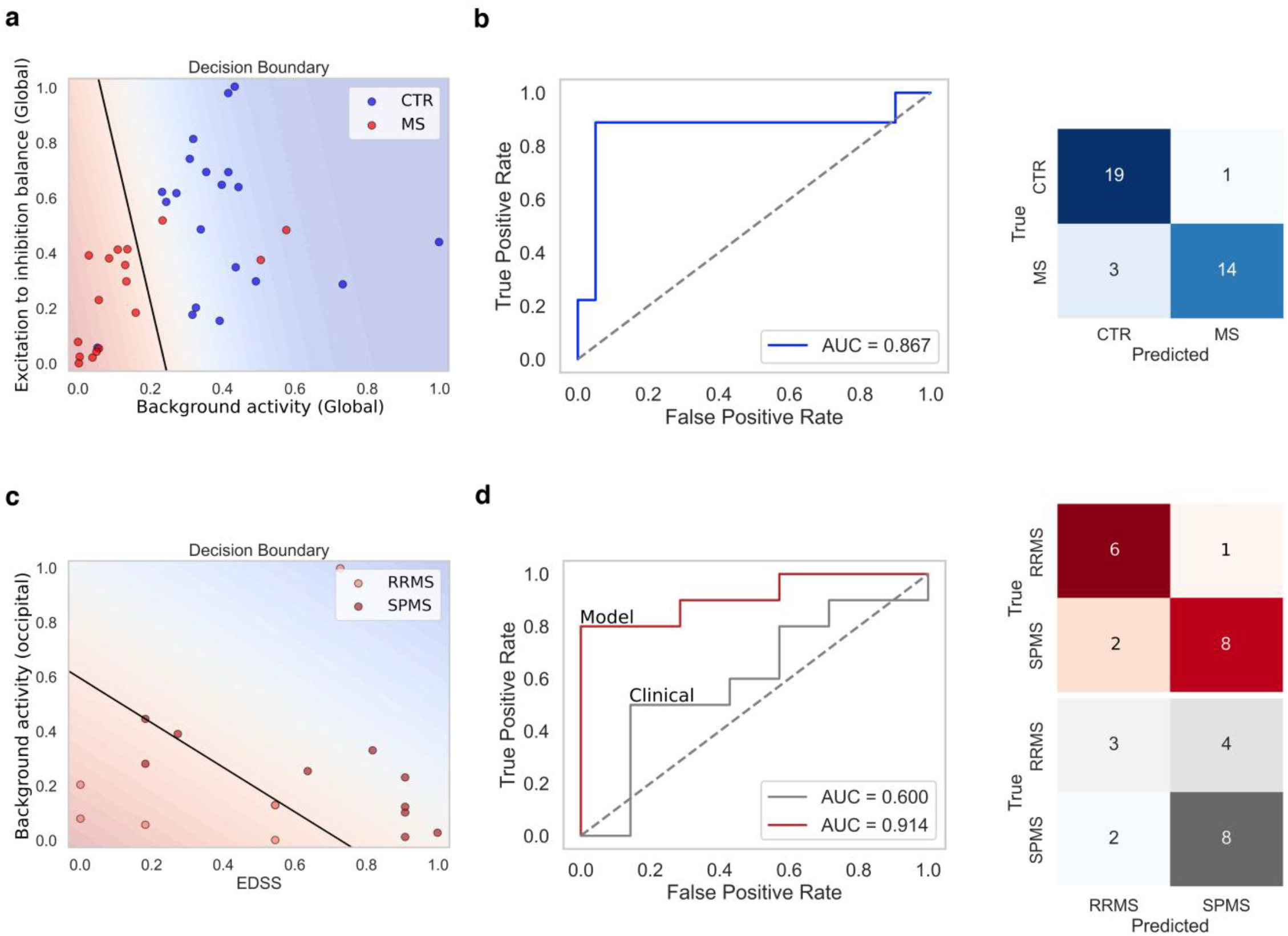
Classification performance of excitability parameters. **(a):** Decision boundary of excitability parameters for the CTR vs. MS classification. **(b):** ROC curve (left) and confusion matrix (right) for the CTR vs. MS classification based on excitability parameters. **(c):** Decision boundary of excitability parameters and clinical features (EDSS) for the classification between MS subtypes: relapsing-remitting (RRMS) and secondary progressive (SPMS). **(d):** ROC curve (left) and confusion matrix (right) for classification between MS subtypes based on excitability parameters (red) and standard clinical features (grey).

We then investigated whether the same framework could improve the differentiation between MS phenotypes (SPMS vs RRMS). The selected model-derived parameters were combined with clinical scales, including EDSS, BDI, FSS, and SDMT, to define two-dimensional decision boundaries (Fig. 2c). While clinical scales alone yielded modest performance (AUC = 0.60, accuracy = 0.61), the combination of EDSS and occipital background activity emerged as the most discriminative feature pair, achieving an AUC of 0.91 and an accuracy of 0.83 (Fig. 2d). These results highlight the added value of model-derived excitability parameters in capturing disease-specific alterations beyond standard clinical measures, allowing for patient stratification both between MS and CTR participants and within the MS groups. Notably, in both the CTR vs. MS and in the SPMS vs. RRMS classifications, model-derived parameters obtained performance that was not achievable using the MEG features used for their derivation (PSD and FC, see Methods). In fact, we tested the MEG features used to derive model parameters as classifying features in the CTR vs. MS classification, obtaining results slightly above chance level (AUC = 0.53, accuracy = 0.58). In the SPMS vs RRMS classification, we tested MEG features alongside clinical scales, following the same procedure adopted for model-derived parameters. In this classification task, the best combination of features was that of EDSS with the alpha-band FC, which obtained AUC = 0.71, accuracy = 0.71, still far lower than the results obtained by model-derived parameters.

### Excitability parameters predict multiple clinical MS scales

We next evaluated the ability of model-derived excitability parameters to predict clinical and cognitive outcomes. Specifically, we compared their predictive performance against a set of conventional predictors, including demographic variables (disease duration, age, sex, and education) combined with the total lesion load derived from MRI.

In contrast, predictions based on standard clinical measures (Fig. 3a) showed substantially lower explanatory power. Specifically, SDMT was predicted with R^2^ = 0.49 (CV R^2^ = 0.53), while markedly weaker performance was observed for BDI (R^2^ = 0.18, CV R^2^ = 0.19), FSS (R^2^ = 0.22, CV R^2^ = 0.23), and EDSS (R^2^ = 0.33, CV R^2^ = 0.35).

**Figure 3:**
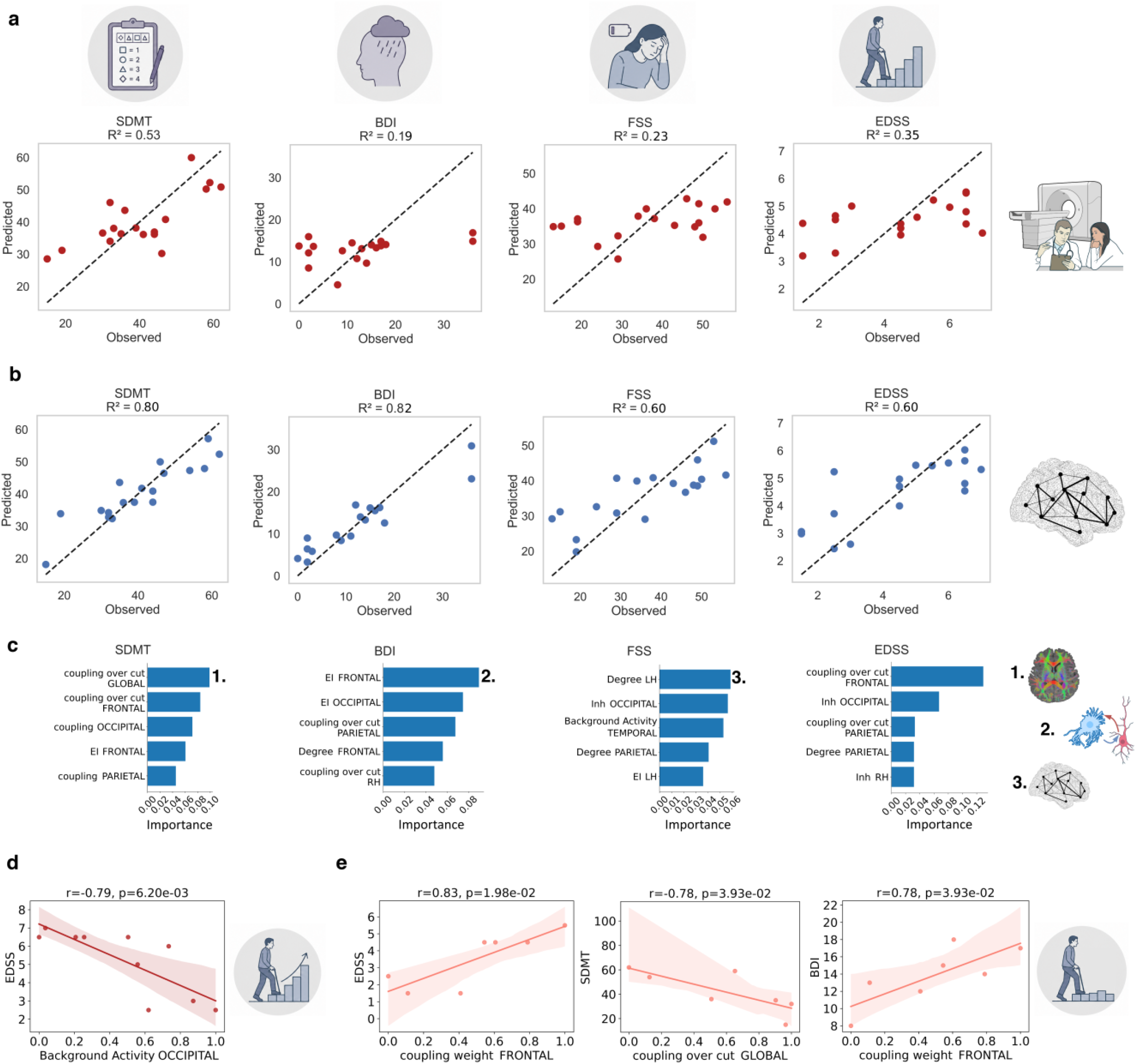
Prediction of MS subscales based on excitability parameters and standard clinical features. **(a):** Prediction of MS subscales (SDMT: Symbol Digit Modalities Test; BDI: Beck Depression Inventory; FSS: Fatigue Severity Scale; EDSS: Expanded Disability Status Scale) based on standard clinical features. **(b):** Prediction of MS subscales based on excitability parameters. **(c):** Importance of excitability parameters for the prediction of different clinical scales. Top predictors were the connectivity coupling over cut (EDSS and SDMT), the excitation to inhibition balance (BDI) and the connective node degree (FSS). **(d):** Predictions of clinical scales in SPMS patients with excitability parameters. **(e):** Statistically significant predictions of clinical scales in RRMS patients using excitability parameters.

Across all scales, excitability parameters consistently obtained superior performance when compared with standard measures. Using model-derived features (Fig. 3b), we observed high predictive performance for cognitive and clinical scores, with R^2^ = 0.77 (CV R^2^ = 0.80) for SDMT and R^2^ = 0.81 (CV R^2^ = 0.82) for BDI. Moderate but robust performance was also observed for FSS (R^2^ = 0.56, CV R^2^ = 0.60) and EDSS (R^2^ = 0.57, CV R^2^ = 0.60).

To further assess the added value of model parameters over standard methods, we also ran a prediction of clinical scales based on the combination of standard clinical measures and MEG features (PSD and FC, see Methods). Crucially, this combined prediction was still far weaker across all clinical scales with respect to that of model-derived parameters: SDMT: R^2^ = 0.58 (CV R^2^ = 0.52), BDI (R^2^ = 0.39, CV R^2^ = 0.40), FSS (R^2^ = 0.23, CV R^2^ = 0.25), and EDSS (R^2^ = 0.33, CV R^2^ = 0.36).

Cross-validated R^2^ values slightly exceeded in-sample estimates in most models. While this pattern may appear counterintuitive, it likely reflects the high variance of individual LOO-CV fold estimates in small samples and should be interpreted with caution. Overall, these results demonstrate that excitability parameters derived from personalized computational models provide substantially greater predictive power for both cognitive and clinical outcomes than traditional demographic and radiological measures.

To further investigate the contribution of individual biomarkers in predicting each clinical scale, we quantified feature importance for each predictive model (Fig. 3c). For SDMT, the most informative features were the global connectivity coupling over cut (importance = 0.099) and frontal connectivity coupling over cut (importance = 0.082). EDSS prediction was primarily driven by frontal connectivity coupling over cut (importance = 0.13) and occipital inhibition (importance = 0.067). For FSS, the strongest contributors were connective node degree in the left hemisphere (importance = 0.058) and occipital inhibition (importance = 0.056). Finally, BDI was best explained by the excitation-to-inhibition balance in the frontal (importance = 0.091) and occipital (importance = 0.076) regions.

We additionally explored subgroup-specific associations between individual biomarkers and clinical scores in RRMS and SPMS patients. In the SPMS subgroup, EDSS was strongly associated with occipital background activity levels (r = -0.79, p = 0.0062; Fig. 3d). In RRMS patients (Fig. 3e), EDSS was significantly predicted by frontal connectivity coupling over cut (r = 0.83, p = 0.020), SDMT by global connectivity coupling over cut (r = -0.78, p = 0.039), and BDI by frontal connectivity coupling (r = -0.78, p = 0.039). These subgroup-specific relationships suggest that distinct excitability parameters may capture different pathological mechanisms underlying clinical impairment across MS phenotypes.

## Discussion

The present study demonstrates that personalized computational models of whole-brain neural dynamics can yield excitability parameters that robustly discriminate between MS patients and healthy controls, stratify MS subtypes, and predict clinical outcomes across multiple domains. These findings extend prior computational neuroimaging work and support the potential of model-derived biomarkers as sensitive, mechanistically interpretable tools for MS characterization^20,21^.

Among the parameters derived from model inversion, background neural activity and the excitation-to-inhibition balance emerged as the most discriminative features for distinguishing MS patients from healthy controls. This is consistent with a growing body of evidence linking MS pathophysiology to alterations in cortical excitability^16,19,37,38^. The sensitivity of these parameters to group-level differences, with effect sizes substantially exceeding those of conventional lesion-based measures, suggests that they reflect functionally meaningful pathological changes that structural MRI fails to resolve.

The disconnection parameter *sevcut*, derived from individual lesion maps and incorporated into the model to modulate effective connectivity, proved particularly informative in predicting clinical scale scores. This result aligns with a lesion-network framework in which the functional impact of focal structural damage is determined not solely by lesion volume but by the position of lesions within the connectome and the resulting disruption of long-range communication^39,40^. Critically, our approach builds directly on the frameworks proposed by Sorrentino et al.^20^ and Mazzara et al.^21^, who demonstrated that model-derived conduction velocity parameters reconstructed from MEG recordings predict individual EDSS scores more accurately than total lesion load, and provided a principled linear mapping between propagation delays and tract-level structural damage. The present work complements this methodology by incorporating lesion topology into a multi-parameter optimization framework, enabling a richer characterization of the relationship between structural damage and functional impairment.

A particularly noteworthy finding concerns the RRMS subgroup, in which model-derived biomarkers (frontal connectivity coupling over cut intensity) exhibited strong and significant correlations with EDSS, SDMT, and BDI scores. This is clinically meaningful as RRMS patients represent a diagnostically challenging population^41^. RRMS patients, in fact, show no significant differences in total lesion load compared with progressive forms^42,43^. Furthermore, in the RRMS subtype, WM lesions often fail to capture the full spectrum of disability with respect to the SPMS subtype^44^. The ability of model-derived parameters to explain individual variability in this subgroup suggests that excitability biomarkers may offer added sensitivity precisely where conventional tools are least informative.

Notably, in both the classification and prediction of clinical scales, model-derived parameters proved to be more informative than the MEG features used for their derivation. This demonstrates how the use of personalized brain modeling might prove essential to obtain information about the patient’s status that is unachievable with standard methodologies, with profound implications for precision medicine.

An important consideration when interpreting these findings is that the mechanistic meaning of model-derived parameters depends on the assumptions embedded in the model. Similar empirical brain dynamics may, in principle, be reproduced by alternative models invoking different latent parameters and, consequently, different pathophysiological interpretations. For this reason, the choice of model should be guided by prior biological knowledge and by the specific clinical question being addressed. This does not undermine the value of mechanistic modeling but rather emphasizes that the epistemic value of inferred parameters depends on whether the model encodes biologically plausible mechanisms relevant to the disease under study. In this respect, the strong predictive performance observed in secondary progressive MS is particularly informative, as this disease stage is characterized by a greater contribution of neurodegenerative processes and gray matter dysfunction, in addition to inflammatory demyelination. The present findings therefore support the relevance of modeling local excitability alterations as a complementary dimension of MS pathophysiology. Nevertheless, MS remains a multifactorial disorder in which inflammatory demyelination, lesion-driven disconnection, altered conduction, and neurodegeneration may coexist. Future extensions of the multiple sclerosis virtual brain-twin framework should therefore aim to integrate these mechanisms into a unified modeling approach to improve both mechanistic interpretability and patient-level predictive accuracy.

Several limitations should be acknowledged. First, the sample size is relatively small (17 MS patients, 20 controls), which constrains statistical power and the generalizability of the classification and prediction models; replication in larger, independent cohorts is warranted. Second, the cross-sectional design precludes causal inference and does not allow assessment of biomarker sensitivity to disease progression over time. Third, the optimization procedure relies on a heuristic genetic algorithm, which may not guarantee convergence to a globally optimal solution, and the selected parameter sets represent approximate rather than exact solutions^45^. Finally, the computational cost of individualized model fitting currently limits scalability for routine clinical application, though ongoing advances in simulation efficiency may address this constraint in the near future.

Overall, computationally inferred excitability parameters not only outperformed standard MEG-derived features and total lesion load in predicting clinical disability scores, but also achieved superior accuracy in discriminating between healthy controls and patients, demonstrating their added diagnostic value over conventional neurophysiological analysis. These findings provide strong proof of concept for the integration of computational modeling into both clinical practice and research pipelines in multiple sclerosis.

## Data Availability

All data produced in the present study are available upon reasonable request to the authors

## Notes

### Competing Interest Statement

The authors have declared no competing interest.

### Author Declarations

The research was conducted in accordance with the Declaration of Helsinki. A written informed consent was obtained from subjects after explanation. The study was approved by the Ethics Committee ASL-NA1 centro (Prot.n.93C.E./Reg. n.14-17OSS).

## References

1. Dobson R, Giovannoni G. Multiple sclerosis – a review. European Journal of Neurology. 2019;26(1):27–40. doi:10.1111/ene.13819

2. Barkhof F. The clinico-radiological paradox in multiple sclerosis revisited. Current Opinion in Neurology. 2002;15(3):239.

3. Mollison D, Sellar R, Bastin M, et al. The clinico-radiological paradox of cognitive function and MRI burden of white matter lesions in people with multiple sclerosis: A systematic review and meta-analysis. PLOS ONE. 2017;12(5):e0177727. doi:10.1371/journal.pone.0177727

4. Brownlee WJ, Hardy TA, Fazekas F, Miller DH. Diagnosis of multiple sclerosis: progress and challenges. The Lancet. 2017;389(10076):1336–1346. doi:10.1016/S0140-6736(16)30959-X

5. Calabrese M, Poretto V, Favaretto A, et al. Cortical lesion load associates with progression of disability in multiple sclerosis. Brain. 2012;135(10):2952–2961. doi:10.1093/brain/aws246

6. Lucchinetti CF, Popescu BFG, Bunyan RF, et al. Inflammatory Cortical Demyelination in Early Multiple Sclerosis. New England Journal of Medicine. 2011;365(23):2188–2197. doi:10.1056/NEJMoa1100648

7. Lombardo MC, Barresi R, Bilotta E, Gargano F, Pantano P, Sammartino M. Demyelination patterns in a mathematical model of multiple sclerosis. J Math Biol. 2017;75(2):373–417. doi:10.1007/s00285-016-1087-0

8. Pirko I, Lucchinetti CF, Sriram S, Bakshi R. Gray matter involvement in multiple sclerosis. Neurology. 2007;68(9):634–642. doi:10.1212/01.wnl.0000250267.85698.7a

9. Fisniku LK, Chard DT, Jackson JS, et al. Gray matter atrophy is related to long-term disability in multiple sclerosis. Annals of Neurology. 2008;64(3):247–254. doi:10.1002/ana.21423

10. Fisher E, Lee JC, Nakamura K, Rudick RA. Gray matter atrophy in multiple sclerosis: A longitudinal study. Annals of Neurology. 2008;64(3):255–265. doi:10.1002/ana.21436

11. Inglese M, Oesingmann N, Casaccia P, Fleysher L. Progressive Multiple Sclerosis and Gray Matter Pathology: An MRI Perspective. Mount Sinai Journal of Medicine: A Journal of Translational and Personalized Medicine. 2011;78(2):258–267. doi:10.1002/msj.20247

12. Rovaris M, Judica E, Gallo A, et al. Grey matter damage predicts the evolution of primary progressive multiple sclerosis at 5 years. Brain. 2006;129(10):2628–2634. doi:10.1093/brain/awl222

13. Mainero C, Caramia F, Pozzilli C, et al. fMRI evidence of brain reorganization during attention and memory tasks in multiple sclerosis. NeuroImage. 2004;21(3):858–867. doi:10.1016/j.neuroimage.2003.10.004

14. Tewarie P, Hillebrand A, Schoonheim MM, et al. Functional brain network analysis using minimum spanning trees in Multiple Sclerosis: An MEG source-space study. NeuroImage. 2014;88:308–318. doi:10.1016/j.neuroimage.2013.10.022

15. Schlaeger R, D’Souza M, Schindler C, Grize L, Kappos L, Fuhr P. Electrophysiological markers and predictors of the disease course in primary progressive multiple sclerosis. Mult Scler. 2014;20(1):51–56. doi:10.1177/1352458513490543

16. Leocani L, Gonzalez-Rosa JJ, Comi G. Neurophysiological correlates of cognitive disturbances in multiple sclerosis. Neurol Sci. 2010;31(2):249–253. doi:10.1007/s10072-010-0398-y

17. Kail R. The neural noise hypothesis: Evidence from processing speed in adults with multiple sclerosis. Aging, Neuropsychology, and Cognition. 1997;4(3):157–165. doi:10.1080/13825589708256644

18. Caramia MD, Palmieri MG, Desiato MT, et al. Brain excitability changes in the relapsing and remitting phases of multiple sclerosis: a study with transcranial magnetic stimulation. Clinical Neurophysiology. 2004;115(4):956–965. doi:10.1016/j.clinph.2003.11.024

19. Neva JL, Lakhani B, Brown KE, et al. Multiple measures of corticospinal excitability are associated with clinical features of multiple sclerosis. Behavioural Brain Research. 2016;297:187–195. doi:10.1016/j.bbr.2015.10.015

20. Sorrentino P, Pathak A, Ziaeemehr A, et al. The virtual multiple sclerosis patient. iScience. 2024;27(7). doi:10.1016/j.isci.2024.110101

21. Mazzara C, Ziaeemehr A, Troisi Lopez E, et al. Mapping Brain Lesions to Conduction Delays: The Next Step for Personalized Brain Models in Multiple Sclerosis. Human Brain Mapping. 2025;46(7):e70219. doi:10.1002/hbm.70219

22. Martí-Juan G, Sastre-Garriga J, Martinez-Heras E, et al. Using The Virtual Brain to study the relationship between structural and functional connectivity in patients with multiple sclerosis: a multicenter study. Cereb Cortex. 2023;33(12):7322–7334. doi:10.1093/cercor/bhad041

23. Pappalardo F, Rajput AM, Motta S. Computational modeling of brain pathologies: the case of multiple sclerosis. Brief Bioinform. 2018;19(2):318–324. doi:10.1093/bib/bbw123

24. Eriksson M, Andersen O, Runmarker B. Long-term follow up of patients with clinically isolated syndromes, relapsing-remitting and secondary progressive multiple sclerosis. Mult Scler. 2003;9(3):260–274. doi:10.1191/1352458503ms914oa

25. Meyer-Moock S, Feng YS, Maeurer M, Dippel FW, Kohlmann T. Systematic literature review and validity evaluation of the Expanded Disability Status Scale (EDSS) and the Multiple Sclerosis Functional Composite (MSFC) in patients with multiple sclerosis. BMC Neurol. 2014;14(1):58. doi:10.1186/1471-2377-14-58

26. Benedict RH, Fishman I, McClellan MM, Bakshi R, Weinstock-Guttman B. Validity of the Beck Depression Inventory-Fast Screen in multiple sclerosis. Mult Scler. 2003;9(4):393–396. doi:10.1191/1352458503ms902oa

27. Flachenecker P, Kümpfel T, Kallmann B, et al. Fatigue in multiple sclerosis: a comparison of different rating scales and correlation to clinical parameters. Mult Scler. 2002;8(6):523–526. doi:10.1191/1352458502ms839oa

28. Benedict RH, DeLuca J, Phillips G, LaRocca N, Hudson LD, Rudick R. Validity of the Symbol Digit Modalities Test as a cognition performance outcome measure for multiple sclerosis. Mult Scler. 2017;23(5):721–733. doi:10.1177/1352458517690821

29. Thompson AJ, Banwell BL, Barkhof F, et al. Diagnosis of multiple sclerosis: 2017 revisions of the McDonald criteria. Lancet Neurol. 2018;17(2):162–173. doi:10.1016/S1474-4422(17)30470-2

30. Deb K, Pratap A, Agarwal S, Meyarivan T. A fast and elitist multiobjective genetic algorithm: NSGA-II. IEEE Transactions on Evolutionary Computation. 2002;6(2):182–197. doi:10.1109/4235.996017

31. Sanz Leon P, Knock SA, Woodman MM, et al. The Virtual Brain: a simulator of primate brain network dynamics. Frontiers in neuroinformatics. 2013;7:10.

32. Sanz-Leon P, Knock SA, Spiegler A, Jirsa VK. Mathematical framework for large-scale brain network modeling in The Virtual Brain. Neuroimage. 2015;111:385–430.

33. Jansen BH, Rit VG. Electroencephalogram and visual evoked potential generation in a mathematical model of coupled cortical columns. Biological cybernetics. 1995;73(4):357–366.

34. Shu N, Duan Y, Xia M, et al. Disrupted topological organization of structural and functional brain connectomes in clinically isolated syndrome and multiple sclerosis. Sci Rep. 2016;6(1):29383. doi:10.1038/srep29383

35. Haider L, Zrzavy T, Hametner S, et al. The topograpy of demyelination and neurodegeneration in the multiple sclerosis brain. Brain. 2016;139(3):807–815. doi:10.1093/brain/awv398

36. Altmann A, Toloşi L, Sander O, Lengauer T. Permutation importance: a corrected feature importance measure. Bioinformatics. 2010;26(10):1340–1347. doi:10.1093/bioinformatics/btq134

37. Ayache SS, Créange A, Farhat WH, et al. Cortical excitability changes over time in progressive multiple sclerosis. Funct Neurol. 2016;30(4):257–263. doi:10.11138/FNeur/2015.30.4.257

38. Vucic S, Burke T, Lenton K, et al. Cortical dysfunction underlies disability in multiple sclerosis. Mult Scler. 2012;18(4):425–432. doi:10.1177/1352458511424308

39. Krieger SC, Cook K, De Nino S, Fletcher M. The topographical model of multiple sclerosis. Neurology Neuroimmunology & Neuroinflammation. 2016;3(5):e279. doi:10.1212/NXI.0000000000000279

40. Altermatt A, Gaetano L, Magon S, et al. Clinical Correlations of Brain Lesion Location in Multiple Sclerosis: Voxel-Based Analysis of a Large Clinical Trial Dataset. Brain Topogr. 2018;31(5):886–894. doi:10.1007/s10548-018-0652-9

41. Filippi M, Preziosa P, Langdon D, et al. Identifying Progression in Multiple Sclerosis: New Perspectives. Annals of Neurology. 2020;88(3):438–452. doi:10.1002/ana.25808

42. Dalton C, Bodini B, Samson R, et al. Brain lesion location and clinical status 20 years after a diagnosis of clinically isolated syndrome suggestive of multiple sclerosis. Mult Scler. 2012;18(3):322–328. doi:10.1177/1352458511420269

43. Kuchling J, Ramien C, Bozin I, et al. Identical lesion morphology in primary progressive and relapsing–remitting MS –an ultrahigh field MRI study. Mult Scler. 2014;20(14):1866–1871. doi:10.1177/1352458514531084

44. Calabrese M, Rocca MA, Atzori M, et al. A 3-year magnetic resonance imaging study of cortical lesions in relapse-onset multiple sclerosis. Annals of Neurology. 2010;67(3):376–383. doi:10.1002/ana.21906

45. Hashemi M, Ziaeemehr A, Woodman MM, Fousek J, Petkoski S, Jirsa VK. Simulation-based inference on virtual brain models of disorders. Mach Learn: Sci Technol. 2024;5(3):035019. doi:10.1088/2632-2153/ad6230

